# DECREASED BREADTH OF THE ANTIBODY RESPONSE TO THE SPIKE PROTEIN OF SARS-CoV-2 AFTER REPEATED VACCINATION

**DOI:** 10.1101/2021.08.12.21261952

**Authors:** Lydia Horndler, Pilar Delgado, Salvador Romero-Pinedo, Marina Quesada, Ivaylo Balabanov, Rocío Laguna-Goya, Patricia Almendro-Vázquez, Miguel A. Llamas, Manuel Fresno, Estela Paz-Artal, Hisse M. van Santen, Stela Álvarez, Asunción Olmo, Balbino Alarcón

## Abstract

The rapid development of vaccines to prevent infection by SARS-CoV-2 virus causing COVID-19 makes necessary to compare the capacity of the different vaccines in terms of development of a protective humoral response. Here, we have used a highly sensitive and reliable flow cytometry method to measure the titers of antibodies of the IgG1 isotype in blood of healthy volunteers after receiving one or two doses of the vaccines being administered in Spain. We took advantage of the multiplexed capacity of the method to measure simultaneously the reactivity of antibodies with the S protein of the original strain Wuhan and the variants B.1.1.7 (Alpha), B.1.617.2 (Delta) and B.1.617.1 (Kappa). We found significant differences in the titer of anti-S antibodies produced after a first dose of the vaccines ChAdOx1 nCov-19/AstraZeneca, mRNA-1273/Moderna, BNT162b2/Pfizer-BioNTech and Ad26.COV.S/Janssen. Most important, we found a relative reduction in the reactivity of the sera with the Alpha, Delta and Kappa variants, versus the Wuhan one, after the second boosting immunization. These data allow to make a comparison of different vaccines in terms of anti-S antibody generation and cast doubts about the convenience of repeatedly immunizing with the same S protein sequence.

## INTRODUCTION

Vaccines to prevent the worst effects of infections by SARS-CoV-2 virus causing COVID-19 have been rapidly developed. This unprecedented fast development of vaccines has allowed a significant reduction in the number of deaths caused by COVID-19 as well as in the number of patients admitted to the intensive care units of hospitals. In Spain, the immunity reached by the widespread vaccination, and immunity due to previous infections, is likely responsible for the low number of deaths caused by the so-called the fifth-wave by the end of July 2021, in spite reaching a number of reported infections similar to that of previous waves in the fall of 2020 and beginning of year 2021 that caused a much higher number of deaths (https://covid19.who.int/region/euro/country/es). The vaccines that have been more commonly administered in Spain are the mRNA vaccines BNT162b2 (BNT) generated by BioNTech/Pfizer, mRNA-1273 by Moderna, and the adenovirus-based vaccines ChaAdOx1 nCov-19 (ChAd) and Ad26.COV2.S (Ad26) by Oxford University/AstraZeneca and Janssen/Johnsson&Johnsson, respectively. All vaccines were demonstrated to be safe and protective in clinical trials [(2–5)], although reports of rare cases of thrombotic thrombocytopenia associated to both the ChAd and Ad26 vaccines have been published [5,6]. However, the emergence of variants of concern (VOC) of SARS-CoV-2, specifically the Delta variant (B.1.617.2) with a very high transmission capacity(6) is threatening the vaccination strategies in Europe and the USA, given the fact that the efficacy of vaccines against the Delta variant is reduce [8], and fully vaccinated people can become infected and spread the virus efficiently to others [9]. The possibility of giving a third boosting vaccination to immunosuppressed patients to increase the titers of antibodies to the spike (S) protein is being considered [10]. Moreover, the spread of the Delta variant even in fully vaccinated people is making authorities of different countries consider the possibility of administering a third dose to the general population (https://english.elpais.com/society/2021-07-23/everything-pointing-to-need-for-a-third-covid-19-vaccine-dose-says-spanish-health-minister.html). The capacity of the Delta variant to infect vaccinated individuals seems to correlate with its ability to escape neutralization by antibodies produced in response to vaccines(6).

Serological tests are usually established by detecting the presence of viral antigen-specific IgG or IgM in the serum of individuals using recombinant fragments of the S or N proteins and tests based on ELISA or lateral flow assay [11,12]. A disadvantage of those tests is that neutralizing antibodies are not directed against the N protein and that recombinant fragments of the S protein miss the quaternary structure of the protein trimer, which is the native form of the spike protein in the viral envelope. Therefore, part of the neutralizing antibodies directed against the native S trimer could be missed in serological tests based on the expression of recombinant proteins. Recently, we have described a flow cytometry (FC) test that makes use of the cell line of hematopoietic origin Jurkat to express the native S protein of SARS-CoV-2 together with a truncated form of the human EGF receptor (huEGFRt) as a normalizing tool [13]. The method allows the accurate determination of seropositivity and the measurement of antibody titers in sera from post-convalescent patients [13]. Here, we have used this FC method to compare the production of antibodies against the S protein in response to the prime immunization and the booster immunization of the four most common vaccines being used in Spain. In addition, we have used the multiplexed capacity of the method to simultaneously measure the reactivity of sera from vaccinated individuals with the S protein of the original Wuhan isolate versus the Alpha, Delta and Kappa isolates as example of VOC. Our results help to stir the debate about the convenience of a third dose of the same vaccines.

## MATERIALS AND METHODS

### Cells

The human T-cell line Jurkat clone E6-1 was acquired from ATCC (TIB-152) and was maintained in complete RPMI 1640 supplemented with 5% fetal bovine serum (FBS, Sigma) in a 5% CO2 incubator. Human embryonic kidney HEK293T cells (ATCC CRL-3216) were maintained in DMEM supplemented with 10% FBS in a 5% CO2 incubator. All cell lines were routinely tested for the absence of mycoplasma.

### Lentiviral vector and Jurkat cell transduction

The generation of Jurkat-S cells expressing the spike protein of the Wuhan variant has been previously described [(1)]. To express the full-length spike S protein of the Alpha (B.1.1.7), Delta (B.1.617.2) and Kappa (B.1.617.1) SARS-CoV-2 variants in the Jurkat cell line, we used the lentiviral vector based on the epHIV-7 plasmid and the spike S proteins sequences were obtained from the Public Health England database (https://www.gov.uk/coronavirus) and synthesized by the company Eurofins Genomics. Wuhan and Alpha (B.1.1.7) variants were expressed in Jurkat CD3+ cells while Delta (B.1.617.2) and Kappa (B.1.617.1) variants were expressed in Jurkat CD3^−^ cells. The Jurkat CD3^−^ cells were generated in our lab using CRISPR/Cas9 system. The EGFP sequence was obtained from the vector pEGFP-C1 (Addgene). Plasmid constructions were generated by the Gibson Assembly® method.

For transduction, lentiviral-transducing supernatants were produced from transfected packaging HEK-293T cells as described previously [20]. Briefly, lentiviruses were obtained by co-transfecting plasmids pCMV-dR (gag/pol) and pMD2.G (VSV envelope protein), using the JetPEI transfection reagent (Polyplus Transfection). Viral supernatants were obtained after 24 and 48 hours of transfection. Polybrene (8 μg/mL) was added to the viral supernatants prior to transduction of Jurkat cells. A total of 5×10^5^ Jurkat cells were plated in a P24 flat-bottom well in 350 μL of RPMI, and 350 μL of viral supernatant were added. Cells were centrifuged for 90 minutes at 2200 rpm and 32ºC and left in culture for 24 hours. Transduced GFP+ or HuEGFRt+ cells were selected by FACS sorting 48 h later and expanded in culture.

### Human sera

A total of 700 human sera were tested. A first set of 50 sera from Empireo obtained in the year 2020 after the first wave of the COVID-19 pandemics. Serum donors filled in a questionnaire to allow their clinical classification according to the following parameters: <u>Asymptomatic</u>, no symptoms; <u>Mild</u>, 3 or more of the following symptoms: non-productive cough, hyperthermia, headache, odynophagia, dyspnea, asthenia, myalgia, ageusia, anosmia, cutaneous involvement; <u>Moderate</u>, 3 or more of the above symptoms plus gastrointestinal symptoms, or more than 3 of the above for 7 or more days; <u>Moderate-Severe</u>, 3 or more of the above symptoms plus pneumonia; <u>Severe</u>, pneumonia requiring hospitalization and intubation. A second set of 52 serum samples were selected from the study “Immune response dynamics as predictor of COViD.19 disease evolution. Implications for therapeutic decision-making” from the Hospital Universitario La Princesa (HUP) approved by the Research Ethics Committee (no. #4070). A third set of 250 sera from nursery homes and VITRO employees. A fourth set of 40 serum samples were obtained from teachers and employees of a secondary school in Madrid (name not revealed to keep anonymity). Finally, a fifth cohort of 250 serum samples were obtained from capillary blood of volunteers working at the CBMSO in the period of January-July 2021, included in the study “*ACE2 as a biomarker with utility for identification of high risk population for SARS-CoV-2 infection and prognosis of evolution in COVID-19”* approved by the Research Ethics Committee (no. #2352). The ethical approvals for the use of the samples applied to the current study. All participants provided written consent to participate in the study which was performed according to the EU guidelines and following the ethical principles of the Declaration of Helsinki.

### Flow cytometry

For the study of relative binding of antibodies to the Alpha versus the Wuhan variants, a 1:1 mix of Jurkat-S(wuhan) and Jurkat-S(Alpha) cells were incubated for 30 min on ice with 1:50 dilutions of human sera in phosphate-buffered saline (PBS), 1% bovine serum albumin (BSA), 0.02% sodium azide. Cells were spun for 5 min at 900 g and the pellet was resuspended in PBS-BSA buffer and spun to eliminate the excess of antibody. Two additional washes were carried out. The cell pellet was finally resuspended in a 1:200 dilution of mouse anti-human IgG1 Fc-PE (Ref.: 9054-09, Southern Biotech) and a 1:200 dilution of the Brilliant Violet 421™ anti-human EGFR Antibody (Ref.: 352911, Biolegend) in PBS-BSA. Samples were then washed, labeled with the viability dye 7AAD and analyzed on a FACSCanto II flow cytometer (Becton-Dickinson) and the data were analyzed with FlowJo software (BD). For the study of the four variants simultaneously, 1:1:1:1 mix of Jurkat-S(wuhan), Jurkat-S(Alpha) Jurkat-S(Delta) and Jurkat-S(Kappa) cells were incubated as previously described, but adding in a dilution 1:200 of the APC anti-human CD3 antibody (BD Ref.: 317317) to discriminate between CD3+ and CD3^−^ cells.

### Confocal microscopy

Spike protein expression in cell membrane was confirmed by confocal microscopy. A total of 10^6^ cells expressing either of the VOCs were incubated with anti -SARS-CoV-2 (COVID-19) spike antibody (GeneTex Cat No. GTX632604) at a 1:150 dilution in phosphate-buffered saline (PBS), 1% bovine serum albumin (BSA), and 0.02% sodium azide for 30 minutes at room temperature, followed by Alexa 555 Donkey anti-mouse IgG1 (ThermoFisher, Cat No A-31570). Cells were then blocked with a 1:100 dilution of normal mouse serum for 15 minutes. After that, cells were labeled with anti-CD45-PE (BD, Cat No. 555483) at a 1:200 dilution for 30 minutes at room temperature. Samples were allowed to bind to poly-l-lysine-coated glass for 30 min and fixed for 10 min at room temperature with 4% paraformaldehyde. Cell nuclei were stained with 1 μg/ml DAPI in PBS for 5 min, and finally, samples were mounted with Prolong Antifade (Molecular probes). Samples were observed by LSM 710 laser scanning confocal microscope. Image processing was performed using Zen software.

### Statistics

One-way ANOVA tests for column analysis of different groups. All data was analyzed using the GraphPad Prism 7 software. Serum samples were received coded from the providers and the experimentalists were blinded to their nature until all data analysis was finalized. Sample analysis was carried out in duplicate or triplicate and all experiments were repeated a minimum of two times.

## RESULTS

### Comparison of IgG1 anti-S titers in response to the four (ChaAd, Moderna, BNT and Ad26) vaccines

We previously used a lentiviral vector to express the full spike “S” protein of SARS-CoV2 with the original Wuhan-1 sequence, followed by a truncated human EGFR protein (huEGFRt) linked by a T2A self-cleaving sequence in transduced cells [13]. This system allows expression of the two proteins from a monocystronic mRNA. We produced transducing supernatants to express the construct in the human leukemic cell line Jurkat. The S protein was previously tested to be folded in its native form at the plasma membrane of Jurkat cells since it was found to be trimeric by electrophoresis in polyacrylamide gels under native conditions and to be able to promote the formation of syncitia between Jurkat and ACE2+ cells [13]. Taking advantage of the fact that the expression of S protein is coupled to that of huEGFRt, we calculated a ratio of mean fluorescence intensities of antibodies against S versus an antibody against huEGFR as direct estimation of the relative quantity of immunoglobulins against the S protein in sera or other fluids from different donors. A conversion between S/EGFR MFI ratios and international WHO standards in BAU/mL is shown in Fig S1. Using this system, we interrogated what was the relative humoral response, in terms of IgG1 production, to the different vaccines used in Spain from January to August 2021. Since the Ad26.COV2.S (Ad26) vaccine was initially planned to be given in one dose and not in two, prime-boost, as the other three vaccines (BNT162b2 [BNT], mRNA-1273 [Moderna], and ChaAdOx1 nCov-19 (ChAd)], we first compared the titer of antibodies elicited in vaccinated healthy individuals 3-4 weeks after the first dose. We found that ChAd (n=37) induced lower titers of IgG1 antiS antibody than Moderna (n=54) and BNT (n=75) (Fig 1A). However, the titers elicited in response to ChAd and Ad26 (n=11) were not significantly different. Compared to the titer of antibody present in a cohort of patients recovered from SARC-CoV-2 infection between March and May 2020 (https://covid19.who.int/region/euro/country/es) (Patients 2020, n=58, Fig 1A), vaccination with one dose of ChAd elicited significantly lower titers (*P*<0.0001, One-way ANOVA test). By contrast, one dose of either BNT or Moderna induced higher titers of antibody than those present in Patients 2020. Unfortunately, the number of samples in the Ad26 group was insufficient to establish a significant comparison with the Patients 2020 cohort. Altogether, these data suggest that the two RNA vaccines (BNT and Moderna) are more efficient than the two adenovirus-based vaccines (Ad26 and ChAd) at inducing IgG1 anti-S antibodies.

**Fig 1.**
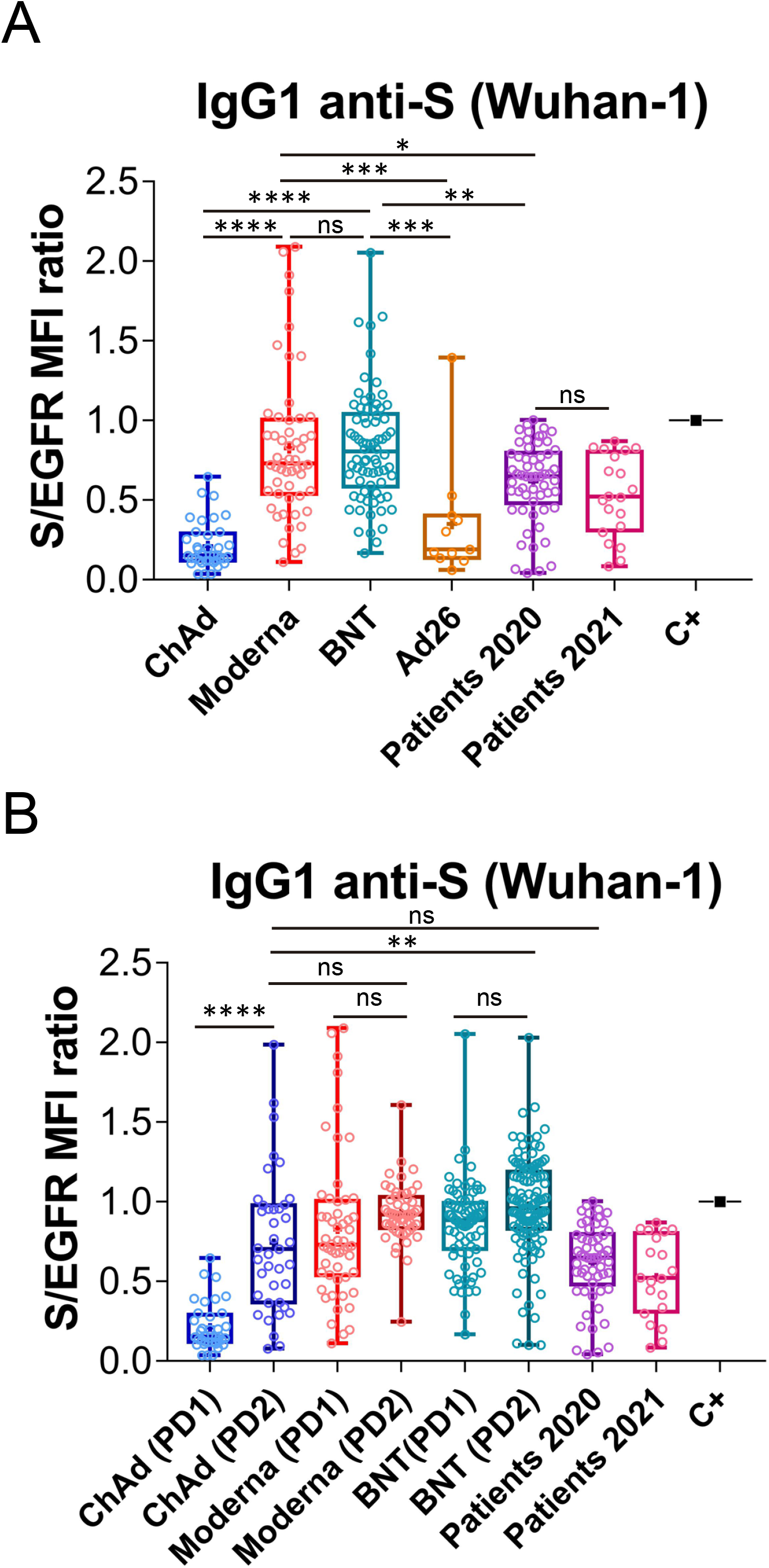
Generation of anti-S protein antibodies in response to the first and second priming-booster doses of different vaccines. **(A)** Generation of antibodies against the S protein of the Wuhan strain 3 weeks (BNT) or 4 weeks (Moderna, ChAd and Ad26) after the first, priming, dose of vaccine to previously uninfected individuals. The ratio of the mean fluorescence intensities (MFI) of anti-S staining and anti-EGFR staining was calculated for each serum sample and normalized to the value of the positive control (C+) sample. All sera were used at a 1:50 dilution. Box and whiskers plots to represent minimum and maximum values as well as the median, and the mean (the mean with a cross) is shown. * *p*<0.05; ** *p*<0.01; *** *p*<0.001; **** *p*<0.0001; ns, not significant (one-way ANOVA test). **(B)** Generation of antibodies against the S protein of the Wuhan variant strain 3 weeks (BNT) or 4 weeks (Moderna, ChAd and Ad26) after the first, priming, dose of vaccine and 4 weeks (all vaccines) after the second, booster dose of vaccine. The ratio mean fluorescence intensities (MFI) of anti-S staining and anti-EGFR staining was calculated for each serum sample and normalized to the value of the positive control (C+) sample. All sera were used at a 1:50 dilution. Box and whiskers plots to represent minimum and maximum values as well as the median, and the mean (the mean with a cross) are shown. The number of samples for each vaccine is shown in the graph. ** *p*<0.01; **** *p*<0.0001; ns, not significant (one-way ANOVA test).

We next compared the titers of anti-S IgG1 generated in response to a priming and a booster dose of the ChAd, Moderna and BNT vaccines. The second dose of ChAd vaccine (PD2; n=39) substantially increased the titer of antibodies to a level comparable to those of Patients 2020 and to those of individuals vaccinated with two doses of Moderna (PD2; n=52; Fig 1B). Nonetheless, the antibody titers elicited by two doses of BNT (PD2, n=119) were still significantly higher than those generated in response to two doses of ChAd (Fig 1B). Interestingly, the titers of IgG1 generated in response to two doses (PD2) of either Moderna or BNT, were not significantly higher than those generated in response to the first priming dose (PD1, Fig 1B). Nonetheless, this result does not argue against other effects of the booster dose of Moderna and BNT vaccines on the humoral response, like improving the memory response or the duration of the protective effect.

### IgG1 titers against the S protein of the Alpha (B.1.1.7) VOC produced after priming-booster vaccination

The flow cytometry Jurkat-S method can be easily multiplexed to measure simultaneously the generation of different antibodies to a single protein or to several proteins [13]. We decided to use this property to measure the generation of antibodies against the S protein of the Wuhan strain in comparison with the antibodies able to bind to the S protein of the Alpha (B.1.1.7) VOC. We thought this comparison might be relevant given that the present vaccines were all generated against the Wuhan strain sequence. The S protein sequence of the Alpha VOC differs in just 7 positions from that of the Wuhan strain (Fig S2). The Alpha VOC became dominant in Spain during the spring of 2021 (https://theconversation.com/the-uk-variant-is-likely-deadlier-more-infectious-and-becoming-dominant-but-the-vaccines-still-work-well-against-it-156951).and therefore, we included in this and previous experiments a cohort of sera from patients recovered from infection by SARS-CoV-2 between March and May 2021 (Patients 2021, n=20, Figs 1 and 2).

**Fig 2.**
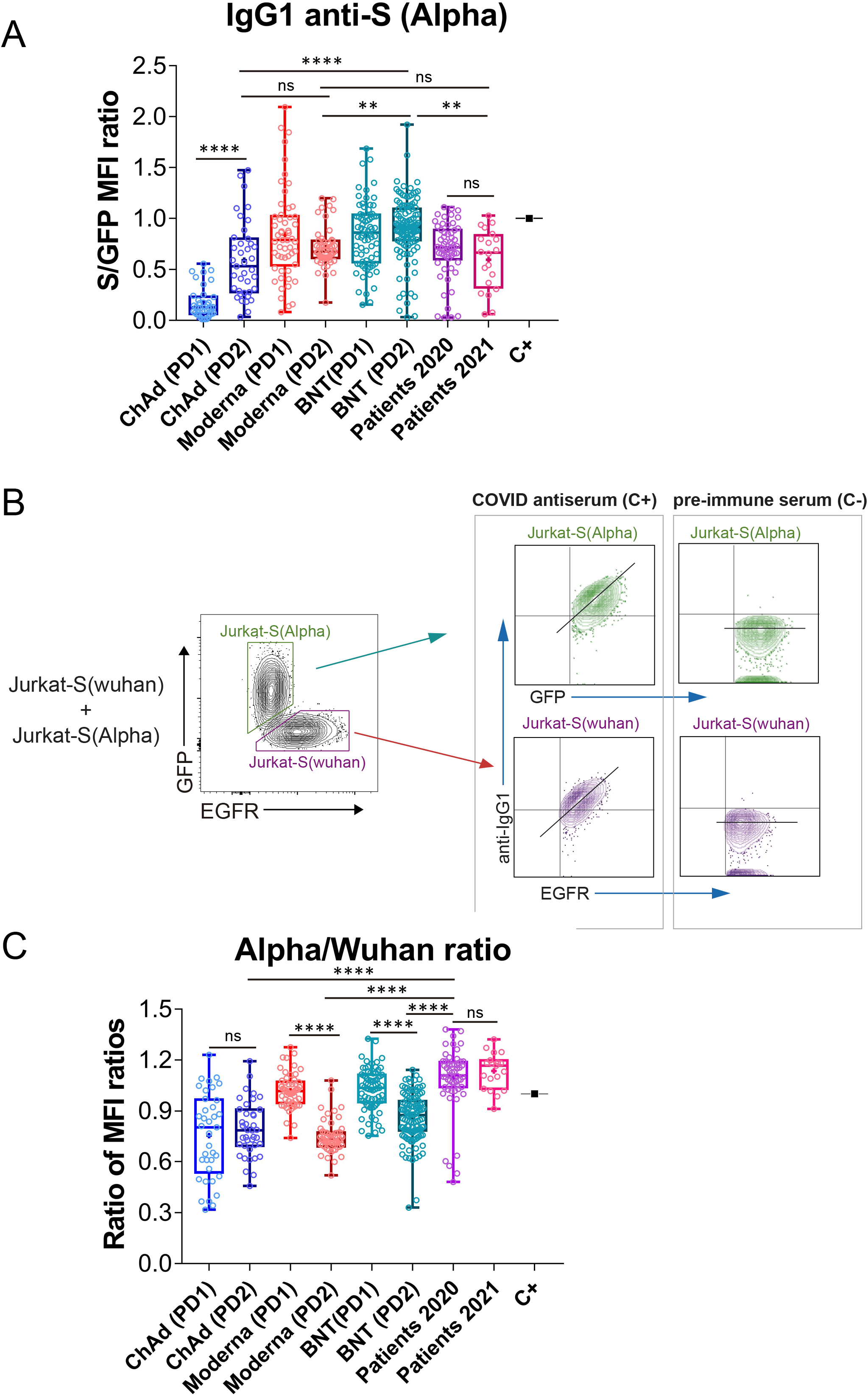
Effect of vaccination on the relative recognition of the Alpha and Wuhan variants by sera of vaccinated individuals. (A) Generation of antibodies against the S protein of the Alpha variant 3 weeks (BNT) or 4 weeks (Moderna, ChAd and Ad26) after the first, priming, dose of vaccine and 4 weeks (all vaccines) after the second, booster dose of vaccine. The ratio mean fluorescence intensities (MFI) of anti-S staining and GFP staining was calculated for each serum sample and normalized to the value of the positive control (C+) sample. All sera were used at a 1:50 dilution. Box and whiskers plots to represent minimum and maximum values as well as the median, and the mean (the mean with a cross) are shown. The number of samples for each vaccine is shown in the graph. ** *p*<0.01; **** *p*<0.0001; ns, not significant (one-way ANOVA test). **(B)** Gating strategy to measure binding to the Wuhan and Alpha variants. Jurkat-S(Wuhan) and Jurkat-S(Alpha) cells were mixed in 1:1 ratio and incubated with either a 1:50 dilution of a positive control serum (C+) from a patient of the first wave in 2020 or a negative control serum (C-) obtained before the COVID-19 pandemics. Cells were subsequently incubated with mouse anti-human IgG1-PE and mouse anti-human EGFR-BV421 antibodies. After incubation, cells were gated according to lymphocyte FSC-A and SSC-A and subsequently on 7-AAD negative cells (7-AAD+ are dead cells). Live cells were subsequently gated on the FL1 channel (for GFP+ cells) and on the FL7 channel (huEGFRt+ cells). Both GFP+ and huEGFRt+ cells were separately analyzed for the fluorescence intensity of anti-IgG1 (S protein, FL2) versus GFP (for Jurkat-S(Alpha)) or for the fluorescence intensity of anti-IgG1 (S protein, FL2) versus EGFR (for Jurkat-S(Wuhan)). A diagonal lane is plotted on the bicolor contour plots corresponding to the C+ staining to indicate a relationship between S protein expression and either GFP or huEGFRt. **(C)** Relative reactivity of the sera tested in (A) against the Alpha and Wuhan variants measured according to the (S/GFP ratio) / (S/EGFR ratio) and plotted individually for each serum donor. **** *p*<0.0001; ns, not significant (one-way ANOVA test).

In order to detect the presence of antibodies against the S protein of the Alpha VOC, we cloned the S sequence of the variant (Fig S2) into the same lentiviral vector used to generate the original Jurkat-S cells [13] but replaced the huEGFRt reporter by EGFP, and generated Jurkat cells stably expressing the S protein of the Alpha variant and EGFP. Using this new Jurkat-S cells (from now on termed Jurkat-S[Alpha]) we could measure the titers of IgG1 antibody generated against the S protein of the Alpha VOC. Our positive control serum from a patient recovered of infection during the first wave of COVID-19 in March 2020 was titrated in both Jurkat-S[Wuhan] and Jurkat-S[Alpha] generating very similar results (Fig S3). We used a non-saturating 1:50 dilution of all sera, as in Fig 1, to compare the response to prime-boost vaccination in terms of generation of IgG1 antibodies capable of binding the Wuhan and Alpha variants simultaneously. We found similar results to those for the Wuhan strain (compare Fig 2A with Fig 1B), namely that the second dose of ChAd significantly increased the titer of antibodies compared to the first dose and that the second dose of Moderna and BNT did not result in a significant increase of IgG1 anti-S titers compared to the first, priming dose (P=0.43 and P=0.92, respectively; One-way ANOVA test).

Taking advantage of the distinct markers used for Jurkat-S(Wuhan) and Jurkat-S(Alpha) cells (EGFR and EGFP, respectively), we could mix equal numbers of both cell lines and incubate the cell mixture with dilutions of sera of patients recovered from infection with SARS-CoV-2 or vaccinated. The analysis by flow cytometry allowed to gate on the GFP+ cells to calculate the ratio of [fluorescence anti-S/GFP fluorescence] as a measurement of the titer of antibodies of the IgG1 isotype against the S protein of the Alpha variant (Fig 2B). Simultaneously, we could gate on the EGFR+ cells to calculate the ratio of [fluorescence anti-S/fluorescence anti-EGFR] as a measurement of the titer of antibodies of the IgG1 isotype against the S protein of the Wuhan strain. The combination of Jurkat-S(Wuhan) and Jurkat-S(Alpha) cells in the same experiment allows to calculate a ratio of ratios (S/GFP vs S/EGFR) that serves to compare sera from vaccinated volunteers in terms of their relative reactivity with the S protein of the Wuhan and Alpha VOC. The mixing of both cell types in this setting would allow the S protein of Alpha to compete with the S protein of Wuhan in order to trap the highest affinity IgG1 antibodies. So, this assay would theoretically serve to detect relative binding to one or other variant by antibodies that have experimented affinity maturation, typical of secondary humoral responses. Interestingly, the second dose of Moderna and the second dose of BNT caused a relative decrease in the reactivity of the antibodies with the Alpha VOC in comparison with the Wuhan-1 isolate (Fig 2C). The second dose of ChAd did not cause either an increase or a decrease in the relative recognition of the S protein of Alpha compared to Wuhan.

A comparison of the relative reactivity of sera from vaccinees with those of people recovered from infection in 2020 and 2021, showed that antibodies produced in response to the second dose of either of the three vaccines were relatively less reactive with the Alpha strain than those produced by natural infection not only in 2021 but also in 2020 (Fig 2C). Those results suggest that the booster immunization with the second dose of the three vaccines, based on the sequence of the Wuhan strain, might impoverish the reactivity of the antibodies with emergent VOC of SARS-CoV-2.

### General decrease of relative reactivity against VOCs after booster vaccination

To determine if the detected decrease in anti –S (Alpha) relative reactivity after the second dose of vaccines was also holding true for more recent VOCs of SARS-CoV-2, we modified the Jurkat-S system to measure IgG1 binding to four variants: Wuhan, Alpha, Delta (B.1.617.2) and Kappa (B.1.617.1). In order to be able to distinguish Jurkat expressing the S protein of the Delta and Kappa variants from the Wuhan and Alpha ones, using the same lentiviral vectors with the EGFR and EGFP markers, we transduced a Jurkat CD3negative cell line generated in our lab by CRISPR/Cas9 deletion of the CD3ζ subunit of the TCR. Those CD3^−^ Jurkat cells were transduced with the lentiviral vectors co-expressing the S protein from Delta VOC and EGFR or co-expressing the S protein from Kappa VOC and EGFP. Using this strategy, we could mix the four Jurkat-S cell lines to incubated them simultaneously with serum samples and later to measure IgG1 binding to the 4 VOCs using CD3, EGFR and GFP markers (Figure 3). Fig 3, illustrates how the positive control serum and a negative control serum stain the four VOC. A positive serum containing IgG1 binding to any of the 4 VOCs produces a diagonal, whereas a negative serum produces a flat line (Fig 3). The S/EGFR and S/GFP MFI ratios produced by the positive control serum for those two Jurkat-S(Delta) and Jurkat-S(Kappa) variants were reduced in terms of absolute numbers, compared to the numbers generated with the MFI ratios for the Wuhan and Alpha VOCs. The differences in the MFI ratio numbers could be intrinsic to the use of CD3^−^ versus wild type Jurkat cells However, the titration of the positive control serum produced a very similar result for the 4 variants (Fig S3), suggesting that the method is valid to detect antibodies against the 4 variants. As control for correct expression of the 4 variants we carried out confocal microscopy analysis of the distribution of the S protein. This showed that the S protein of the 4 variants was expressed at the plasma membrane (Fig S4).

**Fig 3.**
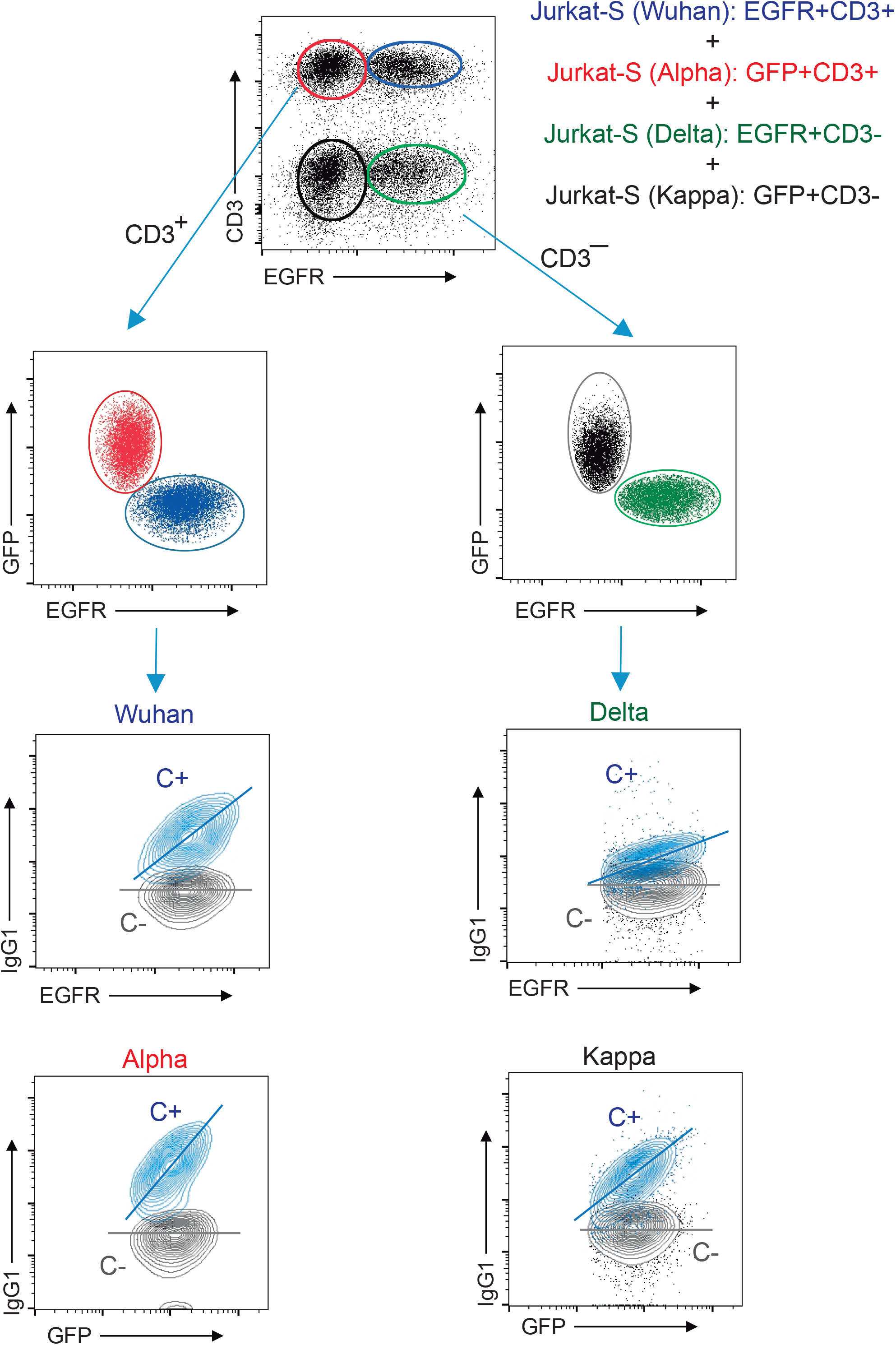
Multiplexed method to measure the reactivity of IgG1 antibodies with the S protein of the Wuhan, Alpha, Delta and Kappa variants. Gating strategy to measure binding to the Wuhan, Alpha, Delta and Kappa variants. Jurkat-S(Wuhan), Jurkat-S(Alpha), Jurkat-S(Delta) and Jurkat-S(Kappa) cells were mixed in 1:1:1:1 ratio and incubated with either a 1:50 dilution of a positive control serum (C+) from a patient of the first wave in 2020 or a negative control serum (C-) obtained before the COVID-19 pandemics. Cells were subsequently incubated with mouse anti-human IgG1-PE, mouse anti-CD3-APC and mouse anti-human EGFR-BV421 antibodies. After incubation, cells were gated according to lymphocyte FSC-A and SSC-A and subsequently on 7-AAD negative cells (7-AAD+ are dead cells). Live cells were subsequently gated on the FL1 channel (for CD3+ cells) and on the FL7 channel (huEGFRt+ cells). This allowed to distinguish four populations of cells: EGFR+CD3+ (Jurkat-S[Wuhan], EGFR^−^CD3+ (Jurkat-S[Alpha]), EGFR+CD3^−^ (Jurkat-S[Delta]) and EGFR^−^CD3^−^ (Jurkat-S[Kappa]). The cells were split between CD3+ and CD3^−^ cells and subsequently analyzed according to EGFR and GFP expression (FL7 and FL1 channels). The four cell populations were separately analyzed for the fluorescence intensity of anti-IgG1 (S protein, FL2) versus GFP (for Jurkat-S[Alpha] and Jurkat-S[Kappa]) and for the fluorescence intensity of anti-IgG1 (S protein, FL2) versus EGFR (for Jurkat-S[Wuhan] and Jurkat-S[Delta]). A diagonal lane is plotted on the bicolor contour plots corresponding to the C+ staining to indicate a relationship between S protein expression and either GFP or huEGFRt.

The effect of vaccination on the relative recognition of the S protein of the Alpha, Delta and Kappa VOC versus that of the Wuhan strain was followed in sera from volunteers of nursing homes who had recovered after infection in 2020 and 2021 and were later vaccinated with BNT (Table 1). Serum samples were taken just before vaccination (pre-Vacc, n=46, Figure 4), 15-21 days after the first dose (PD1, n=49) and 4 weeks after the second dose (PD2, n=70) of the BNT vaccine.

**Fig 4.**
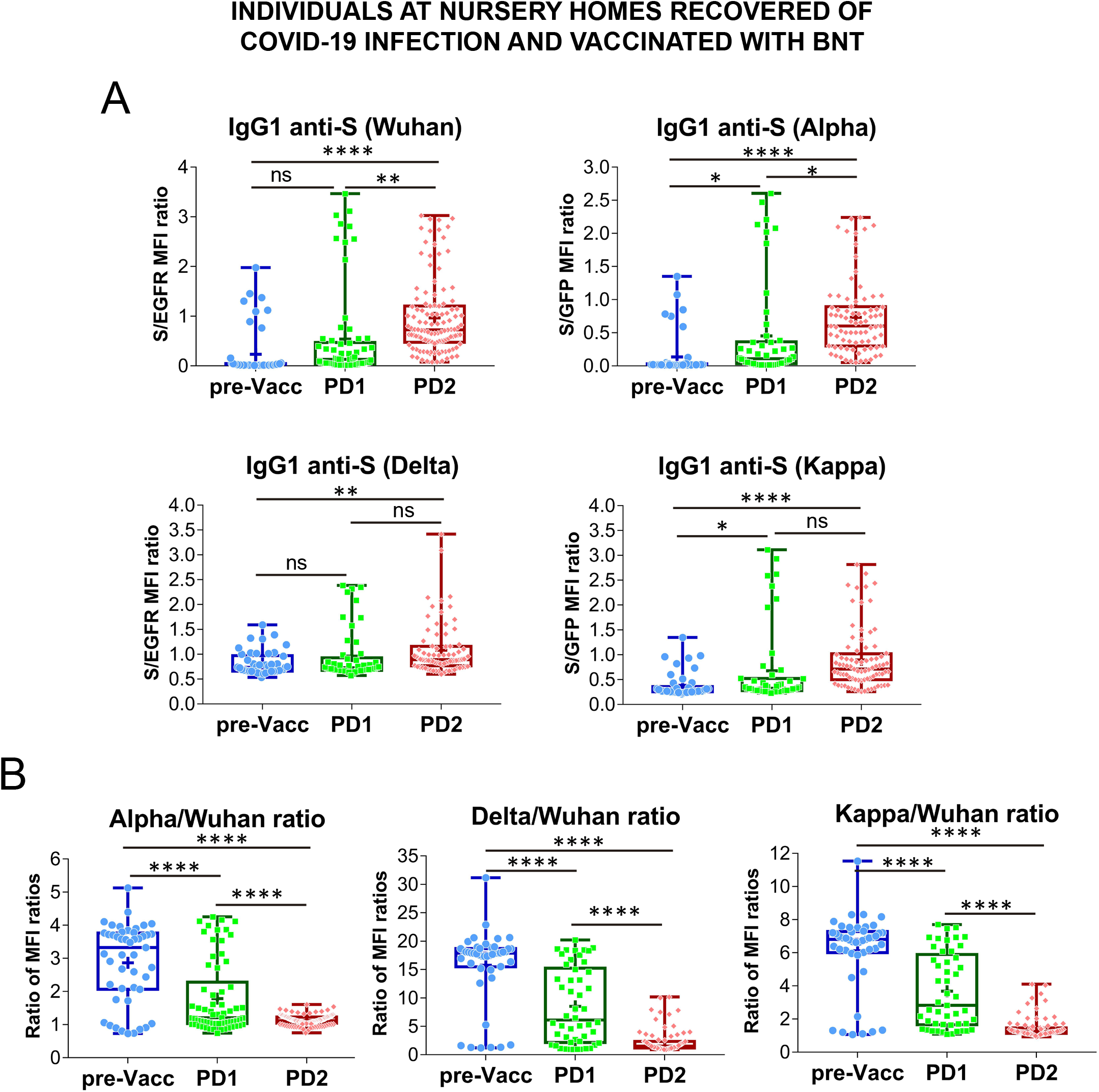
Follow-up of the antibody response to vaccination in nursing homes. **(A)** Generation of antibodies against the S protein of the Wuhan, Alpha, Delta and Kappa variants before (pre-Vacc) and after vaccination with the first dose (PD1) and the second dose (PD2) of BNT/Pfizer. The normalized S/EGFR ratio for reactivity with the Wuhan and Delta variants, and the S/GFP ratio for reactivity with the Alpha and Kappa variants was measured as a quantitative calculation of IgG1 anti-S antibody content. Sera were all used at a 1:50 dilution. Box and whiskers plots to represent minimum and maximum values as well as the median, and the mean (the mean with a cross) are shown. * *p*<0.05; ** *p*<0.01; **** *p*<0.0001 (one-way ANOVA test). **(B)** Relative reactivity of the sera tested in (A) against the Alpha, Delta and Kappa variants versus the Wuhan strain was calculated according to the ratio of S/EGFR or S/GFP ratios and plotted individually for each serum donor. **** *p*<0.0001 (one-way ANOVA test).

The first dose of BNT resulted in a significant increase in antibody reactivity against the Alpha and Kappa VOCs but not against the Wuhan and Delta ones (Fig 4A). Compared to the pre-vaccination samples, the second dose of BNT (PD2) resulted in significantly increased antibody titers against the four VOCs, although the differences between antibody titers induced after PD1 and PD2 were not obvious for the Delta and Kappa ones (Fig 4A). Indeed, the relative reactivity of IgG1 antibodies with the Alpha, Delta and Kappa VOCs compared with that of the Wuhan strain, was significantly reduced after the second dose of BNT compared to the first one (PD1 vs PD2, Fig 4B). These results with samples from vaccinated donors of nursery homes support the Fig 2C data suggesting that the booster vaccination results in a relative loss of recognition of the Alpha, Delta, and Kappa VOC.

## DISCUSSION

In this study we have measured the production of antibodies of the IgG1 isotype to the spike protein of SARS-CoV-2 in response to the four different vaccines most frequently used in Spain. We have compared those data with antibodies present in sera from patients recovered from infection during the first wave of the COVID-19 pandemic in March-June 2020 and with sera from patients recovered from infection during the fourth wave between March-May 2021. Our results show that a priming first dose of the mRNA vaccines BNT/Pfizer and Moderna significantly induced a higher antibody response against the S protein of the Wuhan strain than the two adenovirus-based vaccines ChAd and Ad26, and superior to the titers found in the cohorts of 2020 and 2021 patients. The booster immunization with a second dose did not increase the average titer of antibodies except for the ChAd vaccine. The booster immunization with ChAd resulted in antibody titers that were similar to those generated in response to the booster immunization with Moderna or to the patient cohorts of 2020 and 2021, but still lower than the titers generated in response to the booster immunization with BNT. Therefore, in summary, our results indicate that two doses of the ChAd, BNT and Moderna vaccines are able to induce antibody titers against the original Wuhan strain equal or even superior to those found in post-convalescent patients. However, the message is different when we compared the relative reactivity of the antibodies against the Alpha, Delta and Kappa variants versus the Wuhan strain. Although antibodies made in response to vaccines based on the original Wuhan strain sequence do also bind the other three variants, suggesting that most epitopes are conserved between the vaccine and the variant sequences, there is a relative loss of reactivity with the three VOCs compared to the Wuhan strain occurring upon administration of the booster dose of vaccine. This is somehow expected since repeated immunization with the same antigen sequence leads to the generation of higher affinity antibodies that fit better the epitopes of the immunogen. This increase in affinity has the negative side effect of reducing the “breadth” of the antibodies, that is, their capacity to bind to epitopes that differ slightly from those of the immunogen. Such effect of generating high-affinity antibodies that are however strain-specific has been widely documented in the search of broadly neutralizing antibodies for the human immunodeficiency virus (HIV) [14]. The long-term struggle, and still unsuccessful, to generate a vaccine that prevents infection by the tremendous diversity of clades and mutants of HIV, with the general conclusion that the tested vaccines are effective to prevent infection by the strain used for immunization but not by the myriad of variants found in the field [(8)]. Another example is the result of vaccination with inactivated influenza virus isolated in immunization campaigns, that result in an efficient neutralization of that particular seasonal variant of influenza but results in reduced capacity to neutralize other variants [(9)]. By contrast, antibody responses to natural infection are broad and exhibit different immunodominance patterns(9). Another example of the different breadth in the antibody responses elicited by natural infection versus vaccination is In regard to COVID-19, a broad and sustained polyantigenic immunoreactivity against the S protein and other viral proteins has also been found in COVID-19 patients, in this case associated to the severity of symptoms(10). Our follow-up of the cohort of residents in nursing homes that were vaccinated with two doses of BNT shows that the two doses of vaccine increased the titers of anti-S antibodies against all the four variants. However, compared with the effect of the first dose, the second dose of vaccine did not result in further increases in the antibody titer against the Delta and Kappa variants and had the negative counterpart of reducing the relative reactivity of the antibodies with all three VOCs. This is relevant since the Alpha variant first and the Delta variant later, have been the most rapidly expanding VOCs in Europe and the USA in 2021 (https://www.cdc.gov/coronavirus/2019-ncov/science/science-briefs/fully-vaccinated-people.html). Interestingly, a recent study using an assay similar to the Jurkat-S FC assay but based on the human B cell line Ramos, has also shown that booster vaccination with BNT increases the titer of anti-S antibodies against the Wuhan strain but not against the Alpha and Beta variants (11).

Although our study is limited to measuring the presence of IgG1 against the S protein of SARS-CoV-2, we think our results are relevant to evaluate the overall humoral response, given that IgG1 is the most abundant immunoglobulin in human blood and has potent effector capacities by being able to both fix complement and bind to Fc receptors [19]. Our results showing a loss of relative reactivity with the current VOCs suggest that third doses of the present vaccines, based on the Wuhan sequence, to the general population might not be the best approach to increase the immunity to the emerging VOCs. In our opinion, a third dose should be limited to the population that has been demonstrated to have been poorly responsive to the first two prime and boost doses and not to the general population.

## Data Availability

All data is provided in Table 1 of the manuscript and no deposit in external repositories is required

## Acknowledgments

We are indebted to Valentina Blanco and Tania Gómez for their expert technical assistance. We thank all volunteers of the CBM, school teachers, “Hospital 12 de Octubre” and “Hospital Universitario de la Princesa” for generously participating in the study.

## Author Contributions

LH and PD performed research and analyzed the data; IB helped with experimentation; SR-P, MQ, RL-G, PA-V, MAL, E.-A., SA, and AO provided clinical samples and data and revised the manuscript; MF and HMvS analyzed data and supervised research, BA supervised and designed research, analyzed data and wrote the manuscript.

## DATA AVAILABILITY

This study includes no data deposited in external repositories. Jurkat-S cells are available for academic research upon request to B. Alarcón.

## Supplementary Material

**Fig S1.**
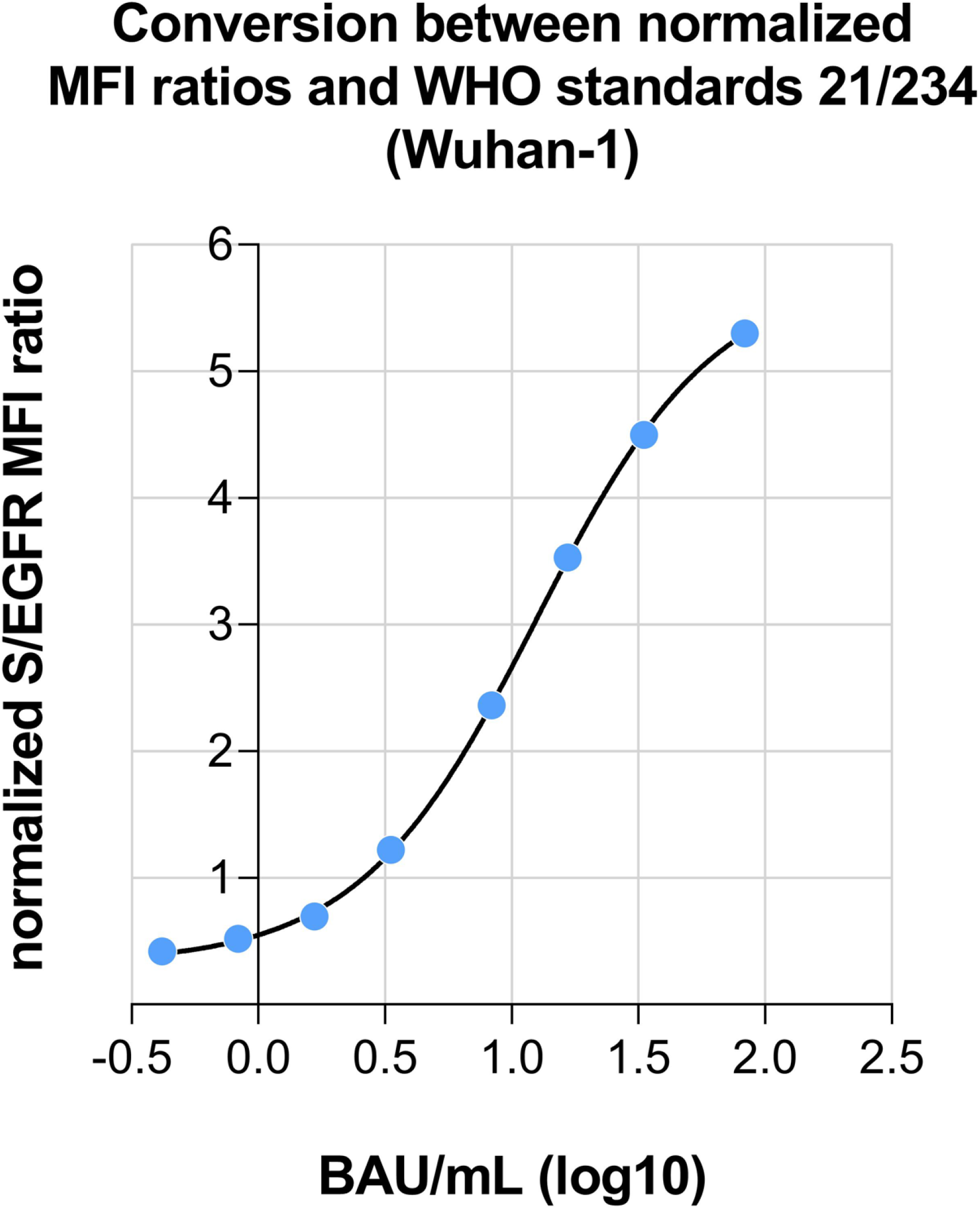
Conversion between normalized ratio and WHO standards 21/234 (Wuhan-1). The international WHO working standard 21/234 was used to obtain a calibration between the normalized S/EGFR MFI ratio on Jurkat-S(Wuhan) cells and Binding Antibody Units (BAU). The datapoints were adjusted to a sigmoidal 4PL fit, where X is in log10 (concentration) with a R^2^= 0.9980.

**Fig S2.**
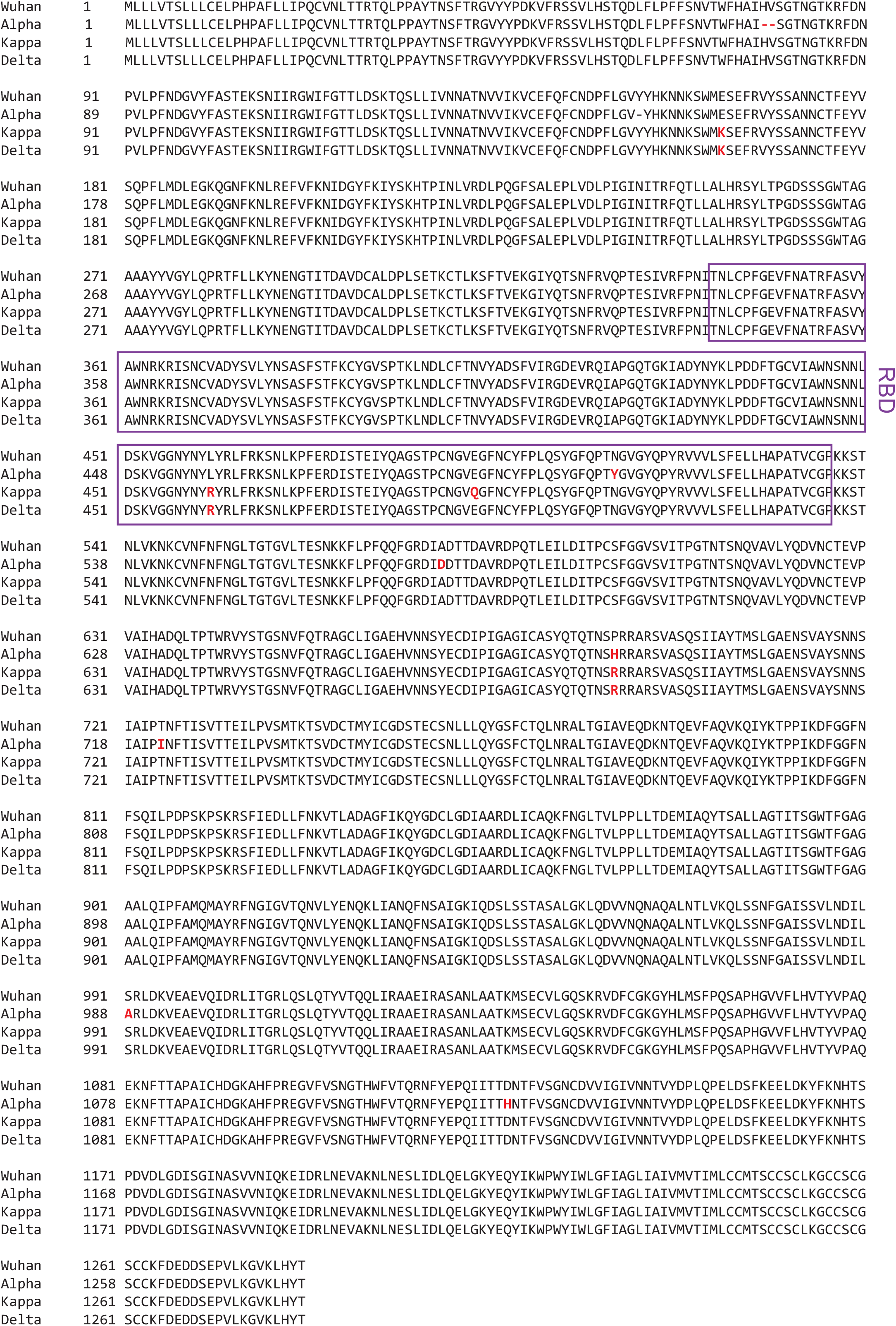
Alignment of the amino acid sequences of the S protein of the Wuhan, Alpha, Delta and Kappa variants of SARS-CoV-2. Amino acid mutations found in the Alpha, Delta or Kappa variants are shown in red bold type. Alpha has 5 amino acid replacements and two amino acid deletions compared to the reference Wuhan strain; Delta has 3 amino acid replacements and Kappa has 4. The sequence corresponding to the Receptor Binding Domain (RBD) is boxed. Alpha has just one amino acid (N to Y) replacement in the RBD, Kappa two (L to R and E to Q) and Delta one (L to R).

**Fig S3.**
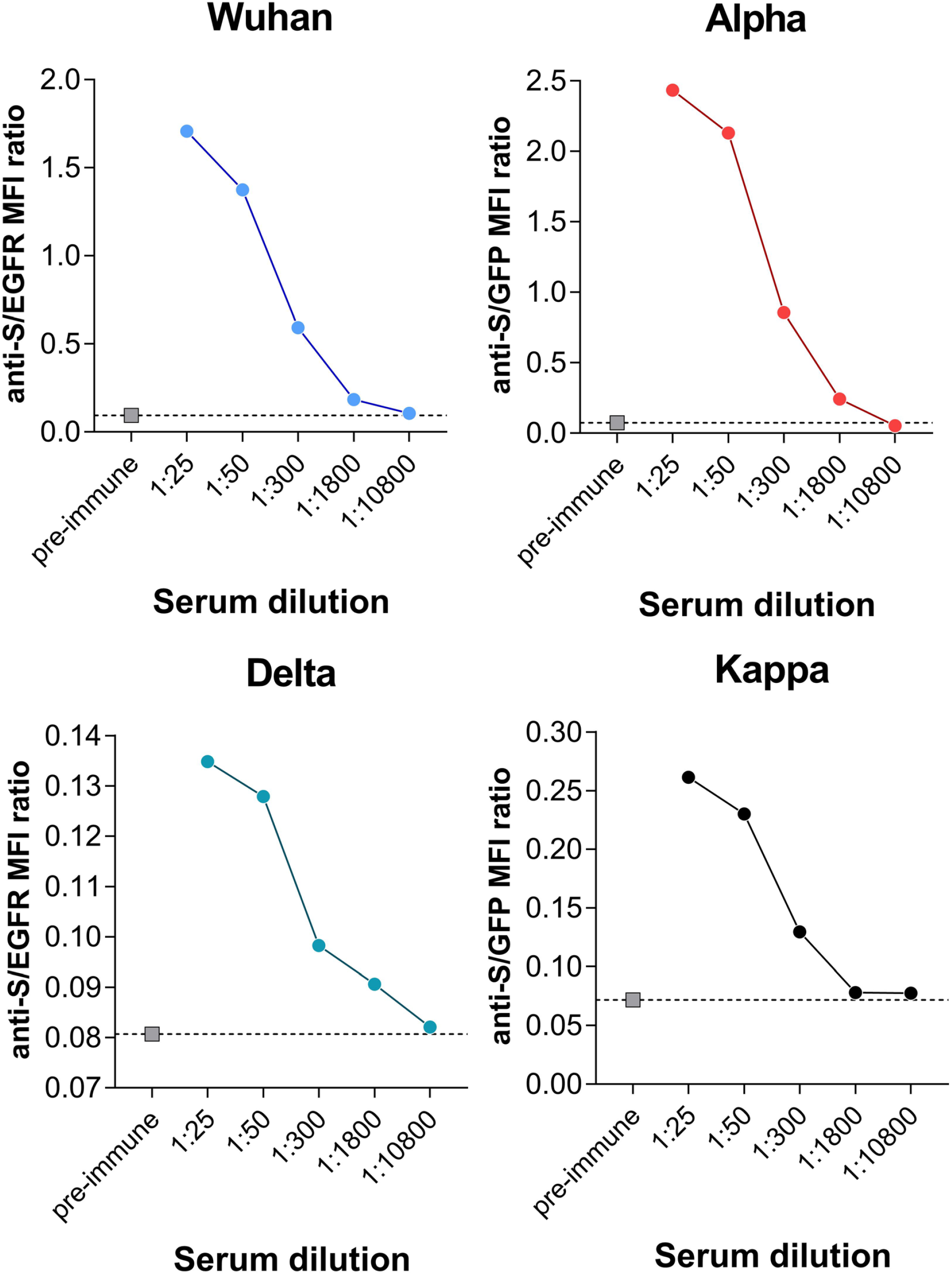
Titration of the different Jurkat-S variant cell lines. Serial dilutions of the positive control serum (C+) used in Fig. 1 and Fig. 2 was tested on the 4 Jurkat-S cell lines to determine their response to decreasing concentrations of antibody. A pre-COVID human serum (dilution 1:50) was used as negative control (grey squares).

**Fig S4.**
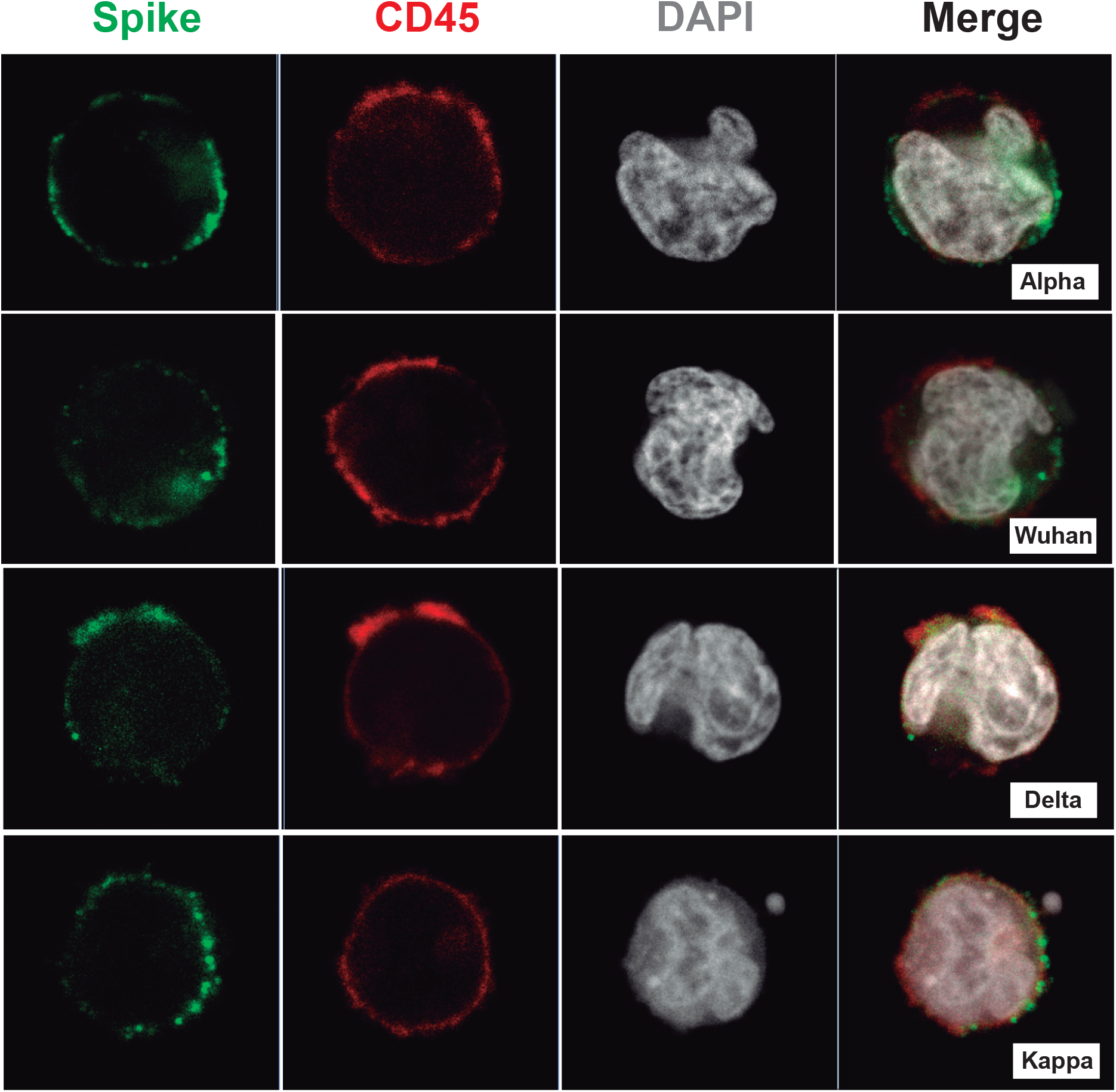
Spike protein expression at the plasma membrane. Mid-plane optical sections of Jurkat-S cells expressing the indicated variants and stained with a mouse monoclonal anti-S protein of SARS-CoV-2 (in green) and a CD45-PE antibody (in red) as a plasma membrane marker. The cell nuclei were stained with DAPI.

**Table S1. Experimental data of Wuhan and Alpha variants**

**Table S2. Experimental data combining Wuhan, Alpha, Delta and Kappa variants**

